# Reliably Assessing Duration of Protection for COVID-19 Vaccines

**DOI:** 10.1101/2021.12.22.21268201

**Authors:** Dan-Yu Lin, Donglin Zeng, Yu Gu, Thomas R Fleming, Philip R Krause

## Abstract

Decision-making about booster dosing for COVID-19 vaccine recipients hinges on reliable methods for evaluating the longevity of vaccine protection. We show that modeling of protection as a piecewise linear function of time since vaccination for the log hazard ratio of the vaccine effect provides more reliable estimates of vaccine effectiveness at the end of an observation period and also more reliably detects plateaus in protective effectiveness as compared with the traditional method of estimating a constant vaccine effect over each time period. This approach will be useful for analyzing data pertaining to COVID-19 vaccines and other vaccines where rapid and reliable understanding of vaccine effectiveness over time is desired.

---

The recent surge of COVID-19 cases has heightened the debate on vaccine boosters, but any decision to give additional doses to those who have been fully vaccinated should be based on careful assessments of the benefits and risks for both individuals and society.^1^ A major consideration in this decision is the durability of vaccine protection against COVID-19 disease, particularly against severe disease. Although evidence on waning vaccine efficacy or effectiveness (VE) is emerging from phase 3 clinical trials and observational studies,^2–8^ more reliable insights could enable properly informed decision-making about the need for and the optimal timing of booster shots. Here, we discuss the sensitivity of existing analyses to waning VE and show how changes in disease risks over calendar time can influence that sensitivity. We then propose an approach with greater sensitivity that can properly adjust for other influencing factors, such as emerging viral variants. Improved assessments of real-world vaccine effectiveness using this approach could also have major implications for decision-making regarding other products.

Waning protection is commonly assessed by comparing VE estimates over successive time periods.^3,5–8^ The VE estimates are obtained under the standard Cox or Poisson model assuming a constant VE over each time period and thus will be referred to as VE_Const_ hereafter. In the presence of waning, VE_Const_ represents an average of the time-varying vaccine effect over the time period, weighted by when the events occur, and thus tends to be higher than the true VE at the end of the time period, especially when the time period is long. In addition, VE_Const_ tends to be less precise when each time period is short, such that larger studies are required to draw firm conclusions.

To obtain more precise and up-to-date estimates of protection, we advocate fitting a Cox model with two time indexes: the event times are measured from the start of the study in calendar time, and the log hazard ratio for the vaccine effect is a continuous, piecewise-linear function of time elapsed since vaccination.^9–10^ The corresponding VE on the hazard rate (VE_HR_) represents exponential deterioration of vaccine effect by time since vaccination.^10–11^ Because it measures the vaccine effect on the instantaneous risk of disease at the current time, rather than an overall benefit over a broad time period, VE_HR_ is more sensitive than VE_Const_ to the level of waning. In addition, measuring time to disease occurrence from trial initiation allows us to account for waxing and waning infection rates and compare disease incidence between the vaccinated and unvaccinated groups at the same calendar time.^9–10,12^

To illustrate the relative sensitivity of these approaches, we simulated a clinical trial mimicking the enrollment pattern of the BNT162b2 study^3^ and the trend of COVID-19 infections occurring in the United States during that trial. We assumed that the true VE_HR_ of a hypothetical vaccine decreases (linearly in the log hazard ratio) from a peak of 95% at full vaccination (i.e., 7 days after dose 2) that lasts one month to 70% at 6 months after full vaccination.^9–10^ The means of VE_Const_ over 1000 replicates are 94.4%, 89.9%, and 81.6% over 0–2, 2–4, and 4–6 months, respectively, which under-represent the degree of waning at the end of each time period (Fig. 1A). The under-estimation by VE_Const_ of the true level of waning was accentuated, even when estimation is performed within two-month intervals, because vaccinations tended to coincide with an early peak in the incidence of infections and then this incidence rate waned for many months thereafter. This resulted in a high percentage of exposures occurring during the earlier part of each two-month interval when the true VE was higher. We simulated a second trial by shifting the enrollment period to 6 months later, such that the period with the strongest vaccine effects coincided with a nadir in exposure rates. Then the means of VE_Const_ over 1000 replicates are 94.7%, 89.5%, and 78.8% over 0–2, 2–4, and 4–6 months, respectively (Fig. 1B). VE_Const_, in essence providing estimates of VE at the mid-points of these 2-month intervals, does not have the same level of overestimation of VE in the second trial relative to the first. In both trials, the estimated VE_HR_ curve is close to the truth (Fig. 1A and 1B).

**Figure 1.**
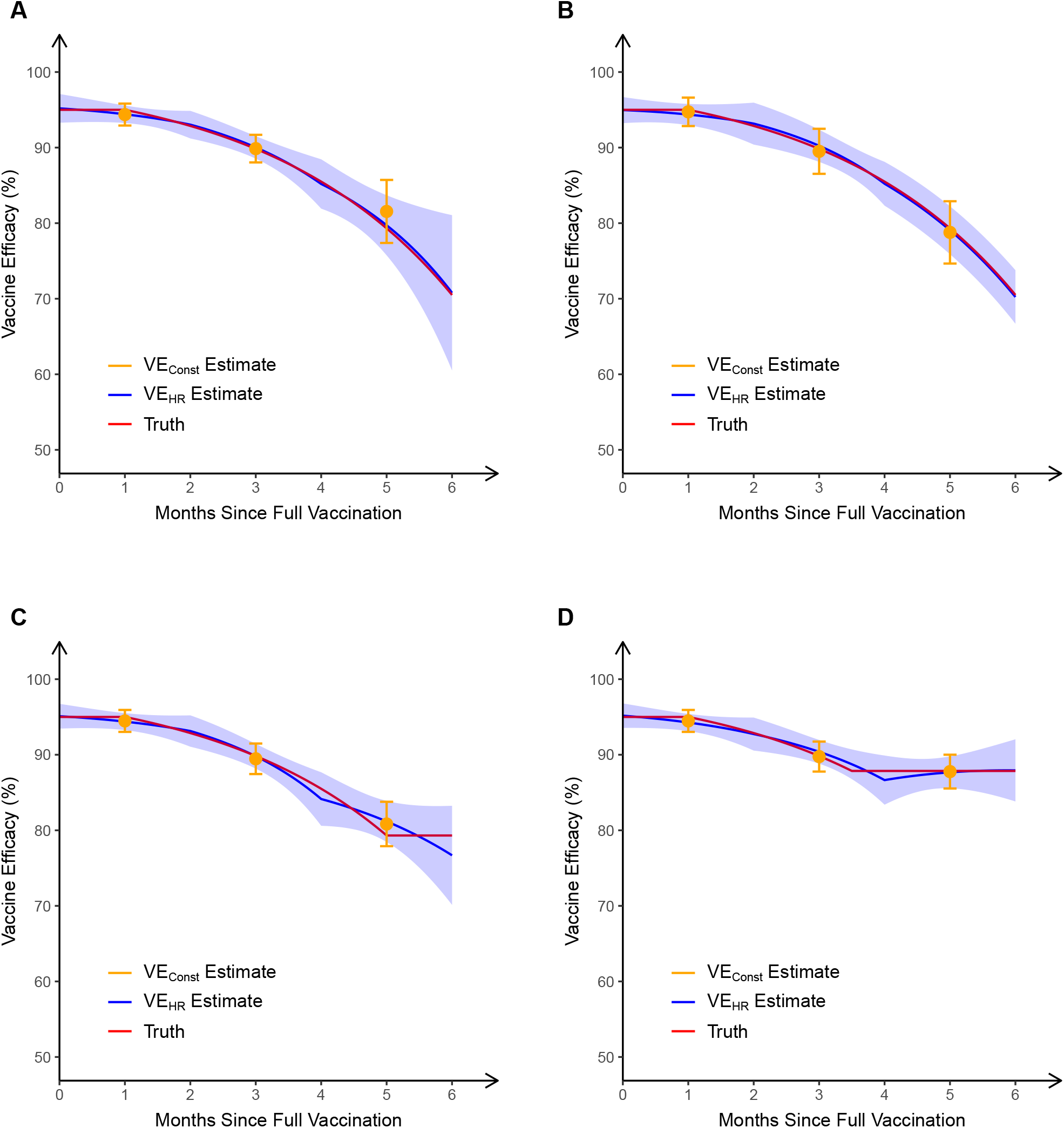
Estimation of vaccine efficacy against symptomatic COVID-19 based on 6 months of follow-up in four simulated clinical trials. In the first two trials, the true VE_HR_ (“truth”) decreases (linearly in the log hazard ratio) from a peak of 95% at full vaccination that lasts one month to 70% at 6 months after full vaccination. In the trial depicted in panel A, most participants received dose 2 at a calendar time coinciding with a peak in infection rates, whereas in the trial depicted in panel B, most participants received dose 2 at a time of low infection rates. In the trials depicted in panels C and D, the true VE_HR_ plateaus at 5 and 3.5 months, respectively. In each trial, VE_Const_ is obtained over 0–2 months, 2–4 months, and 4–6 months post full vaccination, and VE_HR_ is estimated under the Cox model in which the log hazard ratio is a piecewise linear function of time since vaccination, with change points at 0, 2 and 4 months post full vaccination. For each trial, the mean and standard deviation of each estimate over 1000 replicates are shown.

Neutralizing antibodies conferring short-term protection could wane log-linearly, leading to waning of VE over several months yet, for a lengthy duration thereafter, VE could be maintained at a plateau due to cell-mediated or memory immune responses that remain nearly constant over time. Thus, we simulated two more trials by letting the true VE reach a plateau at 5 months post full vaccination in the first trial and at 3.5 months in the second. In the first trial, 6-month VE is somewhat overestimated by VE_Const_ and somewhat underestimated by VE_HR_ (Fig. 1C). In the second trial, both VE_Const_ and VE_HR_ provide estimates of 6-month VE that are close to the truth (Fig. 1D). Importantly, the information obtained from VE_HR_ allows more rapid detection of non-linear changes (such as this plateau) in VE over time, while analysis using VE_Const_ could detect a plateau only with a longer follow-up period.

Due to the crossover of placebo recipients to the vaccine arm, phase 3 trials provide efficacy information only for approximately 6 months post dose 2.^2–3^ Observational studies can provide information about longer-term effectiveness of vaccines. Moreover, large observational databases enable estimation of VE against severe disease and against different viral strains, as well as in various subpopulations. The aforementioned VE_HR_ curve provides similar advantages over VE_Const_ in assessment of waning VE in the observational setting.

The reduction of VE over calendar time^5–6^ or as the time since vaccination increases^7–8^ may be caused by decline of immunity to the primary vaccination, by emergence of new variants that evade antibody recognition, or by both. Comparing VE at a given calendar time among individuals who were vaccinated at different dates allows assessment of waning VE due to declining immunity, and comparing VE at different calendar times for individuals who have been vaccinated for the same amount of time allows assessment of waning VE due to new variants.

The effectiveness of a boosting program, once developed, could be evaluated ideally by large-scale randomization and practically by observational studies. The effect of the booster shot on the hazard rate of disease as a function of time since booster vaccination could be incorporated into the time-varying hazard ratio in the Cox model.^9–10^ The corresponding VE_HR_ curve would provide useful insights into whether and when further boosting is needed.

The proposed approach based on VE_HR_ improves sensitivity for evaluating the true durability of VE using data from phase 3 clinical trials and observational studies, by allowing VE to vary continuously by time post-vaccination and by adjusting for changes in disease incidence over calendar time. Indeed, this approach was recently used in an observational study of COVID-19 vaccine effectiveness in North Carolina.^13^ To reduce confounding bias, analyses of observational data should adjust for individual characteristics (e.g., priority tier, age, sex, race/ethnicity) and the influences of calendar time and geographical location, as well as other factors (e.g., emerging viral variants or vaccination prioritization programs that allow high risk cohorts to be vaccinated first). It is important to recognize that additional factors not easily addressed through modeling (e.g., having a declining number of “controls” over calendar time who remain truly unvaccinated, given an increasing percentage having had infections or vaccinations obtained outside of primary healthcare systems) could be influential. Therefore, changes in underlying immunization rates, disease incidence among vaccinees and controls and follow-up rates among vaccinees and controls over relevant time periods should also be reported.

Properly designed and interpreted analyses regarding durability of VE are invaluable to public health. The more rapid and reliable approach presented here could have implications well beyond COVID-19 vaccine evaluation, since there is increasing interest in using post-marketing effectiveness data to support regulatory and deployment decisions for other vaccines as well. For example, observational studies supported the recent Canadian approval of diphtheria, tetanus, acellular pertussis vaccine administered to pregnant women to protect their infants and long-term observational data supporting effectiveness of a zoster vaccine was included in a US package insert. More rapid availability of results from observational effectiveness studies might thus be useful in confirming effectiveness of vaccines approved under US accelerated approval, EMA conditional approval, or other analogous mechanisms.

## Data Availability

There are no actual data in this manuscript

## Disclaimer

The opinions stated in this article do not necessarily represent those of the US Food and Drug Administration.

